# A genomic epidemiology investigation of yaws re-emergence and bacterial drug resistance selection

**DOI:** 10.1101/2019.12.31.19016220

**Authors:** Mathew A. Beale, Marc Noguera-Julian, Charmie Godornes, Maria Casadellà, Camila González-Beiras, Mariona Parera, Marc Corbacho-Monne, Roger Paredes, Fernando Gonzalez-Candelas, Michael Marks, Sheila A. Lukehart, Nicholas R. Thomson, Oriol Mitjà

## Abstract

**Background:** In a longitudinal study assessing the WHO strategy for yaws eradication using mass azithromycin treatment, we observed resurgence of yaws cases with dominance of a single JG8 sequence type and emergence of azithromycin-resistant *Treponema pallidum*. Here, we analyse genomic changes in the bacterial population using samples collected during the study.

**Methods:** We performed whole bacterial genome sequencing directly on DNA extracted from 37 lesion swabs collected from patients on Lihir Island, Papua New Guinea, between 2013 and 2016. We produced phylogenies and correlated these with temporo-spatial information to investigate the source of new cases and the emergence of five macrolide-resistant cases. We used deep amplicon sequencing of surveillance samples to assess the presence of minority macrolide resistance populations.

**Findings:** We recovered 20 whole *Treponema* genomes, and phylogenetic analysis showed that the re-emerging JG8 sequence type was composed of three bacterial sub-lineages characterised by distinct temporo-spatial patterns. Of five patients with resistant *Treponema*, all epidemiologically linked, we recovered genomes from three and found no variants. Deep sequencing showed that pre-treatment, the index patient harboured fixed macrolide-sensitive *Treponema*, while the post-treatment sample harboured a fixed resistant genotype, as did three of four contact cases. We also found no evidence of pre-existing minority *Treponema* drug-resistance variants in the general population.

**Interpretation:** In this study, re-emergence of yaws cases was polyphyletic, indicating multiple epidemiological sources. However, given the genomic and epidemiological linkage of resistant cases and the rarity of resistance alleles in the general population, it is likely that azithromycin resistance evolved only once in this study, followed by onward dissemination.

**Funding:** Wellcome (Grant #206194), Provintial Deputation of Barcelona (Grant #1940000465).

**Research in context:** *Evidence before this study:* We searched PubMed using the terms “yaws”, “Treponema pallidum”, “mass treatment”, and “azithromycin”, without date or language restrictions. Mass drug administration (MDA) using azithromycin (single-dose 30 mg/kg, maximum 2 g) has been a successful strategy to reduce yaws prevalence in endemic communities, with trials conducted in both Papua New Guinea and Ghana. However, there are no long-term follow-up studies other than in Papua New Guinea, where a resurgence of yaws cases was observed 24-months after MDA. Multilocus sequence typing (MLST) showed a change in *Treponema pallidum* subspecies *pertenue* (TPE) to a single dominant JG8 type, and identified resistant strains with the same MLST type, but this method lacked the resolution to determine the epidemiological mechanism leading to yaws re-emergence and how macrolide resistance manifested.

*Added value of this study:* This study is the first to use whole genome sequencing (WGS) for a detailed investigation of TPE transmission and to apply deep amplicon sequencing to TPE macrolide resistance mutations. Leveraging the increased resolution of WGS over other techniques, we show that re-emergence of yaws after MDA was the result of at least three distinct bacterial sub-lineages with different temporo-spatial properties (likely due to several infected people missing treatment) rather than a single point-source. WGS also shows that patients with macrolide-resistant yaws had genomically indistinguishable TPE, indicating a recent common ancestor, and possibly a recent chain of transmission originating from the index case. Deep amplicon sequencing corroborates that PCR-tested macrolide sensitive samples do not harbour resistant minority populations at detectable levels.

*Implications of all the available evidence:* Our data suggest yaws re-emergence after MDA was driven by multiple sources, and therefore we recommend higher population coverage and intensive post-MDA surveillance. These data show resistance likely evolved once during the trial, rather than being widespread. We recommend careful monitoring of affected communities during post-MDA follow-up in order to rapidly detect new emergence of azithromycin resistance, and consideration for using alternative treatments for cases detected post-MDA.

## Introduction

Yaws, caused by the bacterium *Treponema pallidum* subspecies *pertenue* (TPE), is a neglected tropical disease and a major cause of morbidity in regions of Africa, Asia and the South Pacific^1,2^. Infection with TPE is clinically apparent in the early stages, but population level disease control is complicated by the bacteria’s ability to undergo periods of asymptomatic latency that can span five to ten years, thus evading strategies which target only clinically active cases^1^. This challenge influenced the WHO in designing its Morges Strategy^3^ for yaws eradication using mass drug administration (MDA) of single dose azithromycin for all community members, regardless of infection status.

In a longitudinal study, part of a yaws elimination programme including more than 16,000 residents of Lihir Island, Papua New Guinea, a single MDA with azithromycin reduced the prevalence of yaws from 1·8% to 0·1% after 18 months, but infection began to re-emerge after 24 months, with a significant increase in prevalence to 0·4% at 42 months post-MDA^4^. We previously reported the findings of baseline and follow-up surveys^4^, using a novel multi-locus sequence typing (MLST) scheme^5^ to analyse 239 samples, and showed a transition from a diverse population of TPE sequence types prior to the commencement of MDA to the dominance of a single sequence type (JG8) 24 months after MDA^4^. We further reported the detection of five cases of TPE with genotypic resistance to azithromycin mediated by A2059G mutations in the 23S ribosomal genes, all belonging to the JG8 sequence type^4^.

In this new study, we validated the previous molecular typing system using whole genomes sequenced directly from clinical swab DNA^6^ and used the increased resolution provided by genomic tools in combination with additional epidemiological data to conduct a genomic epidemiology investigation. We aimed to determine the epidemiological mechanisms and source of infection leading to re-emergence of yaws cases after MDA. We also investigated the epidemiological links between cases of drug resistance, and assessed how macrolide resistance emerged under the selection pressure applied by single dose MDA.

## Methods

### Study population

We included all patients with PCR-confirmed TPE during a yaws elimination programme conducted between Apr 1, 2013 and Nov 1, 2016 in Lihir Island, Papua New Guinea, covering a population of over 16,000. All participants, or their parents or guardians, provided oral informed consent to be screened and treated during the study; written informed consent was obtained from all patients with lesions before enrolment. The protocol was approved by the National Medical Research Advisory Committee of the Papua New Guinea Ministry of Health (MRAC number 12·36).

### Data and swabs collection

We matched laboratory results to case reports containing demographic (age, sex, place of origin, place of residence) and clinical data (disease stage, duration). We conducted additional investigations about possible contact with the known drug-resistant cases and sociodemographic risk factors (regular place of socialising, place of religious worship, regular travel to or visitors from another village).

Swabs of lesions from patients with suspected yaws were taken before MDA (April 2013; survey round one), and at six monthly surveys throughout the subsequent 42 months (to October 2016; surveys rounds two to eight), as previously described^4^. Swabs were placed in a lysis/transport buffer that stabilizes DNA and subjected to PCR detection of TPE^7^. Of 777 ulcer samples, 239 were PCR positive for TPE^4^. MLST had been performed previously on all TPE positive samples using a novel three amplicon scheme^5^ and 194 samples were fully-typable. Macrolide resistance single nucleotide polymorphisms (SNPs) were initially detected by restriction fragment length polymorphism (RFLP)^8^.

### Genome sequencing

Of 194 samples from individuals with PCR-confirmed yaws fully-typed by MLST, we selected 50 to provide a representative mix of sequence types, geography, and chronology, and tested TPE pathogen load by qPCR; 32 samples had qPCR C_T_ values below 32 - previous analyses^9^ showed a C_T_ 32 threshold was appropriate for reliable recovery of treponema genomes (Supplementary Figure 1). Five additional samples from patients presenting with genotypic macrolide resistance were selected regardless of C_T_ value. Whole genome sequencing (WGS) was performed directly on TPE DNA extracted from swabs using the pooled sequence-capture approach previously described^6,9^ on Illumina HiSeq 4000 (see Supplementary Methods).

### Phylogenomic analysis of *T. pallidum* infections

We conducted phylogenomic analysis of the study genomes contextualised using 20 publicly available TPE genomes derived from both humans and non-human primates^6,10–14^ (and one *T. pallidum* subspecies *endemicum* genome as outgroup; BosniaA^15^, NZ_CP007548.1). Full methods, conditions and parameters are described in the Supplementary Methods. For genomes without published MLST types, we computationally inferred sequence types from the WGS reads using a custom ARIBA^16^ database containing the published alleles^5^; six recent public genomes contained novel alleles, and we were unable to assign sequence types to these strains. We mapped sequencing reads to a common reference genome (Samoa D; NC_016842.1) to generate a multiple sequence alignment, including genomes where we had sufficient coverage: minimum five reads/site, to call variants for at least 74% of genomic reference positions. We screened our alignment for recombination using Gubbins^17^, and used IQ-Tree^18^ to infer a maximum likelihood phylogeny from the resulting recombination-masked SNP alignment as previously^9^. To further establish the robustness of our tree topology, we performed a Bayesian reanalysis of the recombination-masked SNP alignment, using PhyloBayes^19^ to generate 25,560 trees, and assessed these for alternate edges in a consensus split network and densiTree using the Phangorn^20,21^ package. To assess the phylogenetic patterns underlying clonal expansion of the JG8 sequence type, we performed ancestral reconstruction of the SNPs along the phylogenetic branches within the JG8 lineage and used rPinecone^22^ to define phylogenetically clustered samples as previously described^9^ based on separation by at least 5 SNPs.

### Deep sequencing analysis of minority populations of antibiotic-resistant bacteria

To detect mixed resistance alleles and investigate how the A2059G macrolide resistance mutation first evolved, we designed discriminatory PCR primers specific to the regions flanking the individual copies of 23S rDNA (TPE has two copies of the gene), and generated amplicons for each copy in triplicate for six samples from five patients with resistant strains (see Supplementary Methods). To investigate the broader prevalence of mixed 23S TPE resistance alleles in Lihir patients, we used the same approach to amplify the TPE 23S rDNA from 29 individuals infected with macrolide sensitive TPE. The resulting amplicons were submitted for Illumina sequencing, and the relative proportion of sequencing reads corresponding to each gene copy and each allele (A2058G, A2059G) was analysed.

### Data Availability Statement

Raw sequencing reads from WGS were deposited at the European Nucleotide Archive (ENA) under BioProject PRJEB34799. Raw sequencing reads from deep amplicon sequencing were deposited at the Short Read Archive (SRA) under BioProject PRJNA575636. All accessions used in this project are listed in Supplementary Data 1 and Supplementary Data 2 along with the appropriate sample identifier.

The funders had no role in study design, collection, analysis, or interpretation of data, nor in the writing of the report or the decision to submit the paper for publication.

## Results

Out of 239 patients presenting with PCR positive TPE positive yaws ulcers from the Lihir trial, we performed WGS directly on DNA extracted from clinical swabs for 37. Due to low treponemal load in ulcers, our ability to recover whole genomes directly from clinical samples was restricted even after prefiltering samples according to bacterial load. Including only samples sequenced with a high proportion of genome coverage, we recovered 20 near-whole TPE genomes from the Lihir trial samples (Supplementary Figures 1, 2 and 3).

Of 20 genomes recovered, 16 were of the JG8 sequence type (Figure 1). *In silico* MLST typing of seven closely related TPE genomes from the Solomon Islands^6^ determined that these were also sequence type JG8 under the TPE MLST scheme^5^. In addition, WGS showed a large recombination event previously characterised in the Solomon Island TPE strains^6^, and covering 10 coding sequences (spanning TPESAMD_0856 through to TPESAMD_0866), was present in the JG8 genomes from Lihir as well as the Solomon Islands (Supplementary Figure 4). Phylogenomic analysis of all available TPE genomes showed that the JG8 sequence type is extremely clonal (Figure 1), with all JG8 genomes separated by fewer than 20 pairwise SNPs, and the topology of the JG8 subtree was built on 35 phylogenetically informative sites (Supplementary Figure 4). SNPs were evenly distributed along the genome, but due to low coverage, some of our sample genomes had insufficient sequencing coverage to support all ancestral SNPs (Supplementary Figure 4). However, our phylogenetic reconstruction indicates the most likely placement of genomes. Furthermore, analysis of 25,560 Bayesian trees built using these data found no alternative edges above 1%, showing we have determined the most parsimonious placement (Supplementary Figure 5).

**Figure 1.**
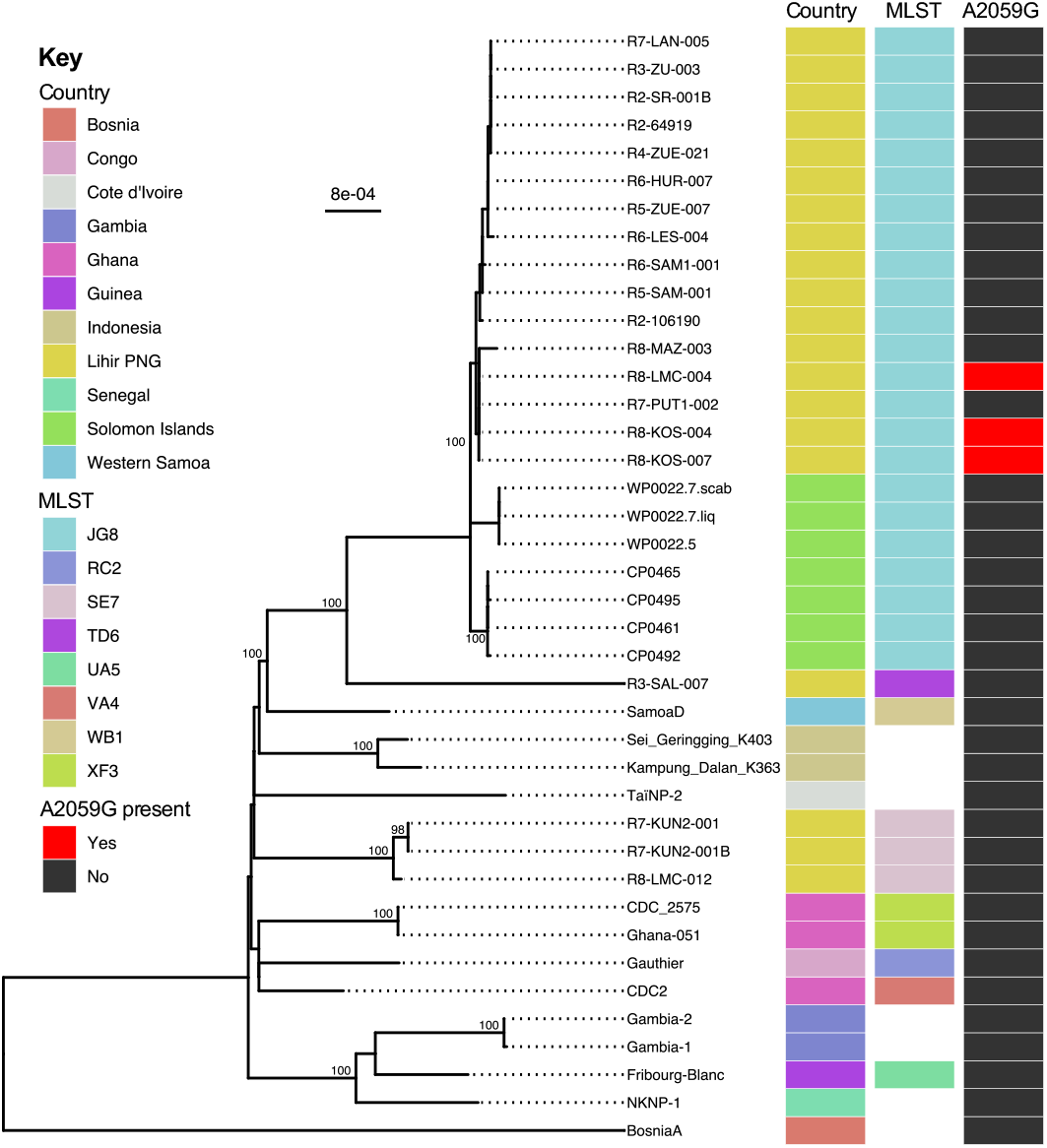
Maximum likelihood WGS phylogeny of 40 TPE genomes, including 20 genomes from Lihir. The JG8 MLST type represents an extremely clonal lineage of TPE found in Lihir and the Solomon Islands, with all JG8 samples separated by ≤ 20 SNPs. Types TD6 and SE7 are genomically distant. Three genomes carrying the A2059G mutation are shown. Branches with Ultra-Fast bootstrap values >=95% are labelled. Branches are scaled by mean nucleotide substitutions/site.

Although the majority (16/20) of Lihir genomes recovered were part of the clonal JG8 sequence type, one genome recovered from a sample taken early in MDA follow-up (R3-SAL-007; Round Three) was of the TD6 sequence type, separated from the JG8 genomes by a long branch of >157 SNPs. Previous analysis of 11 TD6 samples found no history of travel outside of Lihir Island in the preceding six months (TD6 was the second most common sequence type before MDA and after six months, but the proportion of TD6 cases declined thereafter)^4^. Three genomes from Lihir were sequence type SE7, forming a distinct phylogenomic clade more closely related to genomes from Indonesia or Western Samoa than to the JG8 sequence type genomes from Lihir (Figure 1). We previously reported that nine of eleven patients infected with sequence type SE7 reported travel outside of Lihir Island^4^.

Previous MLST analysis showed JG8 was the most common sequence type prior to the MDA, but increased in relative frequency during the MDA follow-up period, and was strongly associated with the re-emergence of yaws cases 24-months after MDA^4^. We investigated whether the spread of the JG8 sequence type could be explained by a single clonal expansion, or through multiple epidemiological routes. Using ancestral reconstruction of the 35 phylogenetically discriminatory SNPs in the JG8 sequence type, we found five distinct phylogenetic clusters (sub-lineages) separated by at least five SNPs, including three sub-lineages from Lihir (Figure 2; sub-lineages JG8.c2, JG8.c3, JG8.c5). Sub-lineage JG8.c2 samples (n=11) were found in five different villages (Hurtol, Landolam, Lissel, Samo, Zuen), whilst sub-lineage JG8.c3 samples (n=5) were found in three villages (Kosmayun, Put Put, Zuen); a sub-lineage JG8.c5 sample from Mazuz village was separated from the JG8.c3 genomes by 6 SNPs. All three JG8 sub-lineages from Lihir were detected in the later rounds of post-MDA follow-up, indicating they were all involved in yaws re-emergence. As Figure 2 shows, rather than being the result of a single clonal expansion, the cases had distinct geographical, temporal and phylogenomic properties, reflecting separate epidemiological and evolutionary histories.

**Figure 2.**
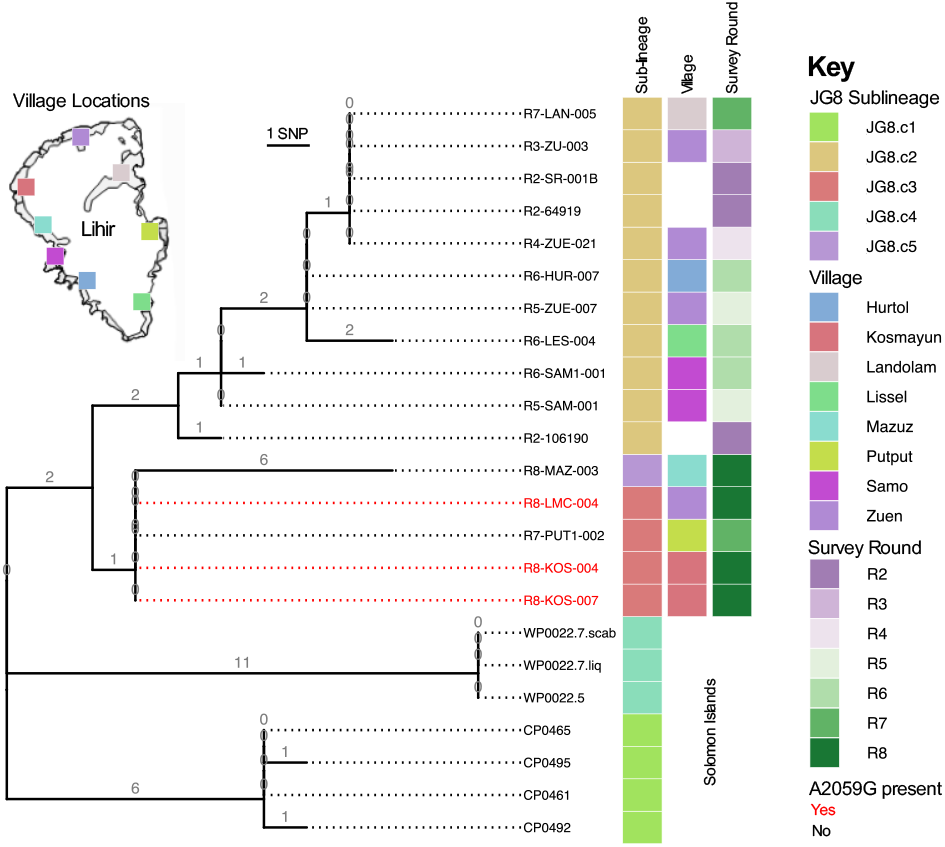
JG8 subtree phylogeny shows clustering of samples by phylogenetic sub-lineage, survey round, village, as well as macrolide resistance. Phylogeny includes 16 genomes from Lihir and 7 from the Solomon Islands. Coloured tracks show phylogenomic sub-lineage, village and survey round number. Village data were unavailable for three patient samples (in white). Branches are scaled and labelled according to the number of non-recombining SNPs. Tip labels for macrolide resistant strains are in red.

We intentionally oversampled and examined a putative transmission network of five cases with treatment failure^4^; all of the TPE samples from these cases were of the JG8 sequence type, and all carried the A2059G mutation linked to azithromycin resistance in *Treponema pallidum* subspecies *pallidum* (TPA)^8^. We identified Patient 1 (sample R7-KOS-003) as representing the earliest macrolide resistant case (index case; May 2016), with the patient reporting having an ulcer for at least three months before formal diagnosis. Nine months earlier (October 2015), Patient 1 had been diagnosed and treated with azithromycin for yaws; RFLP testing showed that this earlier infection was macrolide sensitive (A2059). Epidemiological tracing identified potential transmission links between all five resistant cases (Figure 3) – four of the children were from the same village (Kosmayun), and the index case (Patient 1) was a sibling of Patient 2, playmate of Patients 3 and 4, and school friends with Patient 5 (sample R8-LMC-004). This link between Patients 1 and 5 was not described previously^4^, and was discovered only during follow-up interview; Patient 5 is from Zuen village, 7 kilometres from Kosmayun, but both attend a school in Sale, equidistant between the villages. Two further cases from Kosmayun (Patients 6 and 7) epidemiologically linked to Patient 4 (sibling and playmate) were identified within the timeframe of the investigation; both were infected with macrolide sensitive (A2059) TPE.

**Figure 3.**
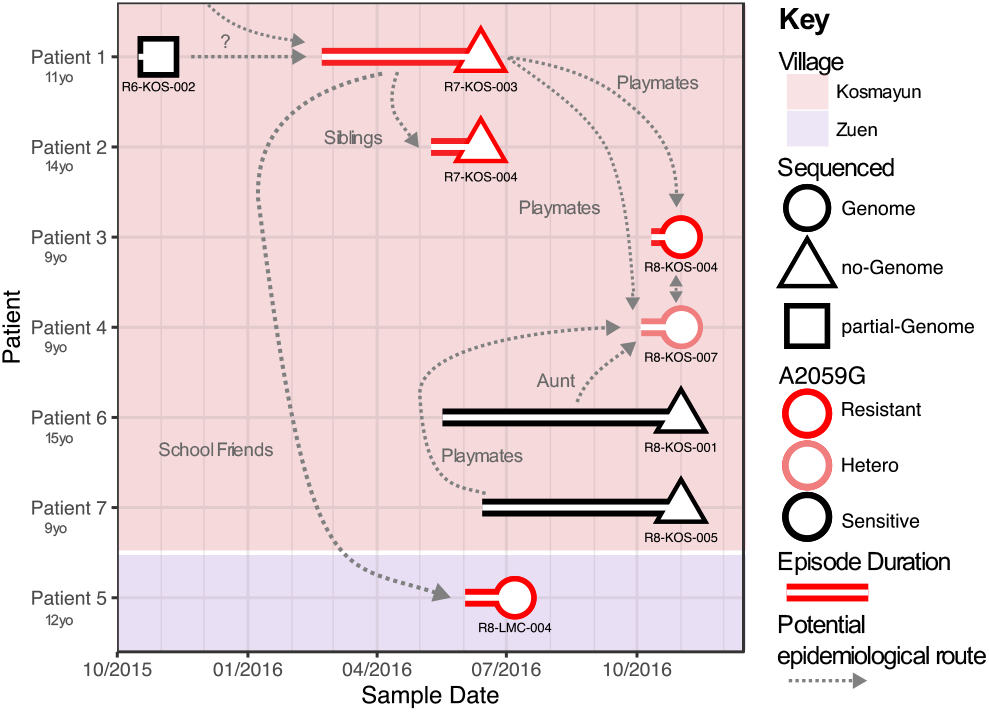
Epidemiological relationships between macrolide resistant samples. Epidemiological relationships between five macrolide resistant samples. Episode duration prior to sampling is based on patient reporting. Patient 1, living in Kosmayun village, was positive for wildtype TPE in October 2015, but follow-up in June 2016 identified A2059G resistant TPE in Patient 1 and sibling Patient 2. A third patient (Patient 5) was identified in Zuen village in July 2016 (school mate of Patient 1). Two more patients from Kosmayun (playmates of Patient 1) were detected with resistant TPE in November 2016. All patient samples were MLST typed as JG8, and of three near-whole genomes recovered from three patients, all were identical over the length recovered. A partial genome (54% of length covered) was obtained from the index case (Patient 1, R6-KOS-002), and this was identical to the others over the common regions.

We were able to recover near-complete TPE genomes from three samples with close epidemiologically relevant contacts (Patients 3, 4 and 5). We compared the 35 discriminatory sites from these three genomes (R8-KOS-004, R8-LMC-004 and R8-KOS-007); in three-way pairwise comparisons we identified no SNPs. However due to insufficient coverage, five or six sites in each comparison had to be masked and excluded from the analysis. This meant we could not rule out the possibility of a SNP being present at those positions. The three genomes were otherwise identical. However, another JG8 genome (R7-PUT1-002) was also identical to the other three samples over the 35 sites (apart from 3-5 ‘n’ sites), but this sample lacked either an epidemiological link to the other cases, or the resistance allele (variants in the 23S region, including A2059G, were excluded from pairwise counts) – these four genomes together comprise sub-lineage JG8.c3 in Figure 2. We attempted to sequence TPE from the pre-treatment October 2015 Patient 1 sample (R6-KOS-002; lacking the macrolide resistance allele), but due to low pathogen load we were only able to recover 54% of the genome; this was identical to R8-KOS-007 and R8-KOS-004 over the high quality common recovered regions. These findings imply that the resistant cases share a recent common ancestor.

We used deep amplicon sequencing (Table 1, Supplementary Data 2) to investigate the evolution of TPE drug-resistance variants in Patient 1. For the earlier wildtype (A2059) sample, we obtained over 8000x sequencing coverage for each 23S rRNA allele, at each gene copy, with three technical replicates. A maximum of 18 reads (<0·22% A2059G) supported the A2059G mutation in any one replicate – well below our threshold for minority variant calling (1%) and equivalent to background sequencing error. This suggests that either the mutation was present but below our limit of detection, that the mutation appeared subsequent to this sample being taken, or this represented a case of reinfection, transmitted from an unsampled individual. Deep sequencing from Patient’s 1 post-treatment sample as well as three of four contact cases with A2059G identified using RFLP, showed a fixed genotype, with >99% of sequencing reads (from a minimum of 797 reads) carrying the A2059G mutation at both 23S copies. In Patient 4 (sample R8-KOS-007), we found only 91% (Table 1; range 87-95% over three technical replicates, consistent for both gene copies) of reads corresponded to A2059G. Manual examination of the WGS reads (derived from a different DNA extract of the same swab) showed a mixture of correctly paired and mapped reads with both alleles at position A2059, consistent with a heterozygous SNP (i.e. a mixed population of sensitive and resistant bacteria). We had insufficient coverage from the WGS to robustly probe genome-wide minority variants and exclude the possibility of coinfection in this patient.

**Table 1.**
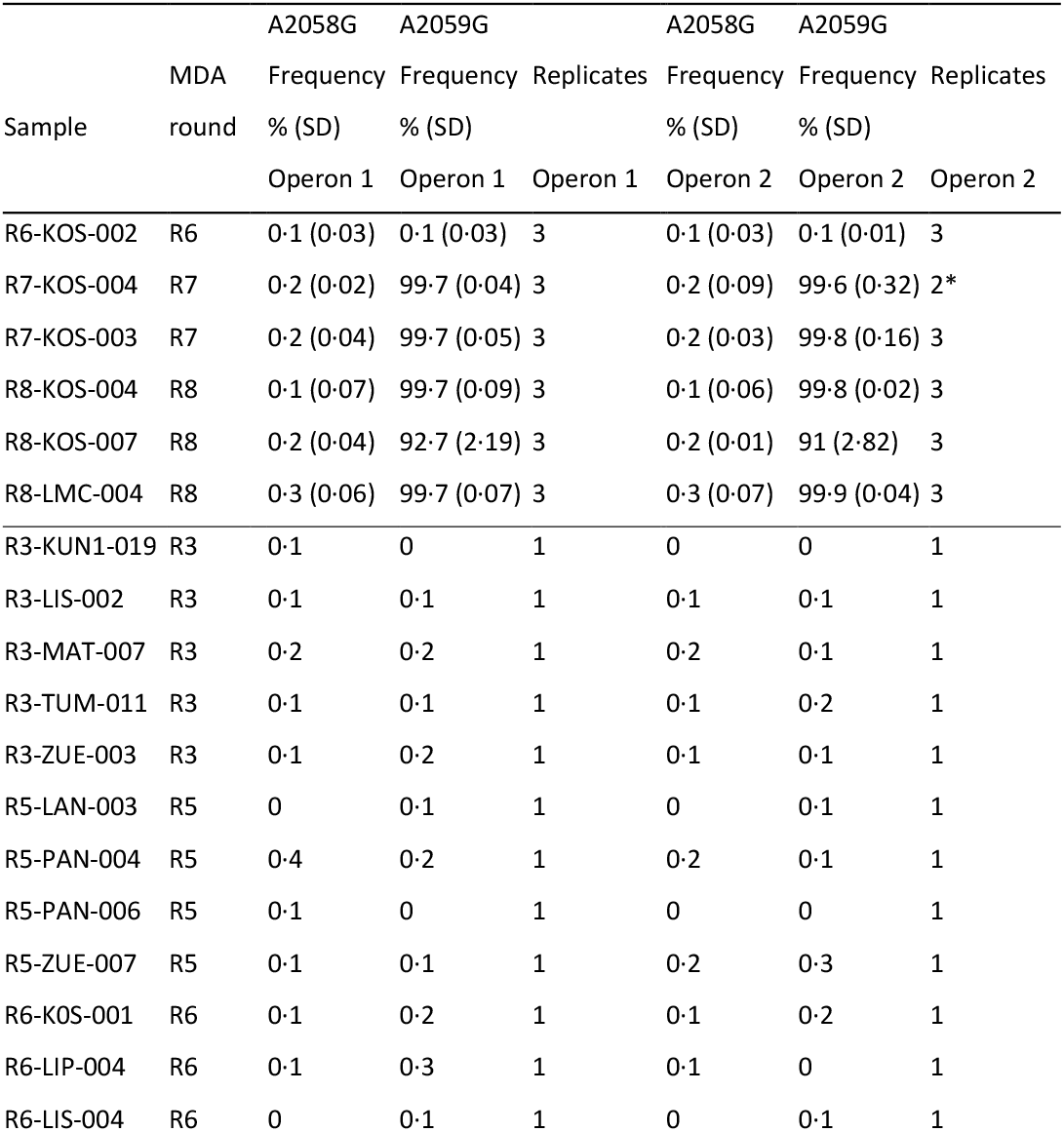
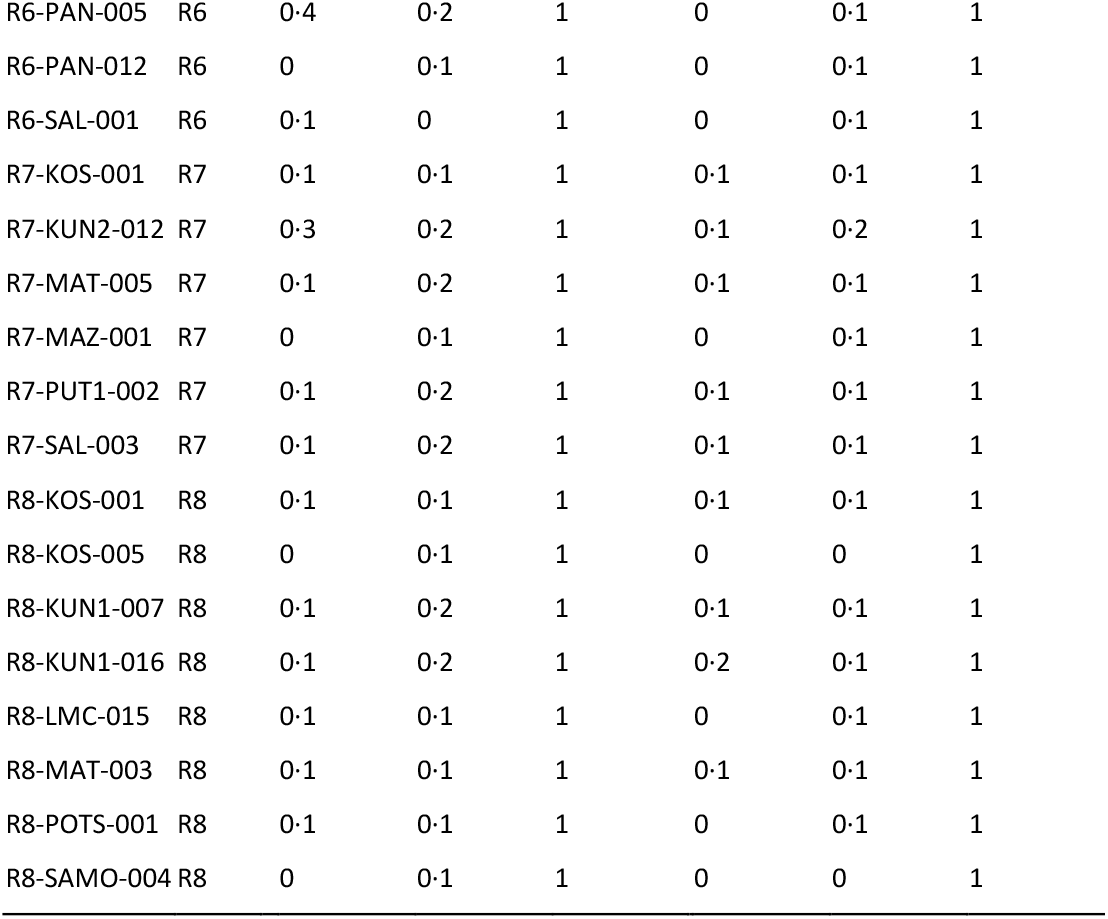
Deep 23S amplicon sequencing of 35 yaws swabs, showing % reads supporting ‘G’ allele at positions A2058 and A2059 for 23S operons 1 and 2. All samples met the minimum threshold of 1000 reads required for inclusion. Samples with RFLP positive A2059G results were tested in triplicate, and for these the Standard Deviation (SD) is shown. *Sample R7KOS004 was tested in triplicate, but one operon 2 reaction failed to return sufficient reads.

To investigate the prevalence of minority TPE drug-resistance variants more broadly, twenty-nine samples were randomly selected from throughout the post-MDA surveillance period (Supplementary Figures 2 and 3) from participants residing in 15 of 28 villages on Lihir Island, nine female and twenty male, median age (IQR) 9 (7-12) years, with four of six MLST types represented. Using the same deep amplicon sequencing approach (but with a single technical replicate), we found no evidence for low frequency macrolide resistance at either position A2058G or A2059G in any of these samples. The finding suggests that the five cases in which A2059G was detected were the result of an isolated *de novo* evolutionary event, consistent with the epidemiological and phylogenomic evidence that these cases are all linked.

## Discussion

Building upon our previous work in which we resolved yaws molecular diagnostic failures through genomics^6^, here we use the increased resolution afforded by WGS phylogenomic analyses to deconvolute the problem of post-MDA yaws re-emergence and macrolide resistance. Single-dose mass treatment with azithromycin has proven to be a valuable tool in low and middle income countries, directed towards the elimination of both trachoma^23,24^ and of yaws^25,26^. In the context of yaws eradication efforts, re-emergence of cases after treatment and bacterial drug resistance represent critical checkpoints for this strategy.

Although a small proportion of cases identified during follow-up were of genetically distinct MLST types (e.g. SE7), consistent with importation of strains from outside Lihir Island, the majority of re-emergent cases were of the endemic JG8 sequence type. The MDA intervention had a lasting effect on the TPE population structure in Lihir, with a diverse TPE population coming to be replaced by a clonal expansion of the JG8 sequence type, the most common lineage prior to the MDA. Using high resolution WGS, we show that the re-emerging JG8 samples are split into at least three distinct phylogenetic sub-lineages. Contextualised by our previous estimate that the mean molecular clock rate of *T. pallidum* is approximately one SNP every four to five years^9^, the geographical dispersal and genetic separation of samples such as R8-MAZ-003 in sub-lineage JG8.c5 from other samples by six SNPs is indicative of distinctly different evolutionary histories within the JG8 sequence type. These sub-lineages support clinical observations that MDA treatment escape is related to multiple epidemiological routes rather than a single point source (either local or imported), consistent with multiple cases of missed MDA, followed by reactivation of latent TPE.

We further describe the genomic relatedness of TPE among the first five people diagnosed with macrolide resistant yaws, representing 2·4 % of post-MDA cases. Macrolide resistance in *T. pallidum* is thought to be caused by either of two mutations targeting the two genomic copies of the TPE 23S gene: A2058G has been linked to multiple independent evolutionary events in TPA^9^, whilst A2059G (the only variant detected in this study) is less common. In this study, all individuals with resistant TPE were epidemiologically linked, had the same A2059G mutation, and JG8.c3 genomes sequenced from three cases were identical over the supported sites. Given the thoroughness of the surveillance and RFLP resistance testing in the post-MDA study population, it is unlikely that we failed to identify additional resistant TPE cases. The most parsimonious interpretation of these data is that a single *de novo* evolutionary mutation conferred macrolide resistance, followed by onward transmission. An alternative explanation for the cluster of macrolide resistant TPE described in this study would be independent A2059G (but not A2058G) resistance mutations arising in five separate patients infected with genomically identical TPE, with the epidemiological linkage being coincidental.

Interestingly, we did not detect the A2059G mutation in the index case (Patient 1) prior to treatment, and this patient’s initial lesion resolved after treatment with azithromycin, suggesting the mutation was acquired after commencement of treatment. This could either be due to selection of *ab initio* resistance in the original strain, or through reinfection from an unsampled individual during the latter months of the treatment phase when the drug levels in tissue were waning. Such a scenario of reinfection during the sub-therapeutic period might explain selection of a resistant subpopulation.

Independent analyses of the sample from Patient 4 indicated that this patient’s TPE was heterozygous for the macrolide resistance allele in both copies of 23S rRNA (91% A2059G). Assuming this epidemiologically-linked patient was indeed part of the same transmission network (rather than a second *de novo* evolutionary event or transmission from an unsampled resistant case), we envisage two possible explanations for this heterozygosity. Firstly, the patient was coinfected with a second wildtype JG8 TPE strain; we were unable to exclude the possibility of coinfection through minority variant analysis of the WGS reads. Of note, Patient 4 shared close epidemiological links with Patients 6 and 7, both infected with wildtype JG8 sequence type TPE, making coinfection a plausible scenario. Secondly, that the patient’s TPE A2059G allele had acquired a sequential mutation back to wildtype. During this study we identified samples with a qPCR C_T_ as low as 25·9 (3652 copies/μl; Supplementary Figure 1, Supplementary Data 1), obtained from a DNA extraction of 100 μl. Ulcer swabs can therefore contain >300,000 bacterial TPE copies, and this represents a small subsample of the bacteria present in a yaws ulcer. We therefore consider it likely that, given time, such a mutation to wildtype A2059 could have occurred in a lesion simply by chance. However, for any mutation to reach a frequency of 9% in a lesion (as detected in Patient 4) implies selective pressure (e.g. fitness cost). We had insufficient data to conclusively discriminate between these two scenarios, but consider coinfection to be a more likely scenario.

Although it appears likely that resistance evolved only once in this study, there is clearly a risk of further emergence. Our broader sampling and deep sequencing from patients throughout the survey period showed that, in this study, TPE samples sensitive according to RFLP methods did not harbour A2058G or A2059G mutations even at low frequency. Whilst these mutations might occur naturally within a patient at low frequency, we would expect them to be selectively neutral or slightly detrimental, and to disappear within a few generations without the selective pressure of drug exposure to drive them to fixation. It will be important in future work to characterise appropriate thresholds for when a minority resistance allele is likely to evolve into a resistant case after drug exposure, as has been done in the HIV field^27^. However, recent analysis of global syphilis lineages suggests that once macrolide resistance evolves and is selected for in a patient, it can persist in lineages^9^ without any evidence of a fitness cost to possessing such variants. Broader MDA with azithromycin may well therefore promote resistance in TPE, and consideration should therefore be given to alternative treatment options for patients demonstrating new or persistent yaws infections post-MDA.

Our study has several strengths. First, we combined detailed epidemiological analysis of temporo-spatial data and phylogenomic reconstruction of bacterial genomes resulting in the best evidence available to understand yaws transmission and bacterial population evolution after MDA. Using genomics enabled previously unachievable levels of strain discrimination, allowing us to determine that resurgent JG8 cases originated from multiple sources rather than a single point source. We recovered 20 near-complete TPE genomes, more than doubling the number of published genomes. However, the inherent limitations of high contamination and low bacterial load in yaws swabs prevented us from sampling the population more completely, such that we may have missed unsampled diversity. Second, we recovered near-complete TPE genomes from three macrolide resistant cases, and moving beyond coarse MLST-level groupings, we show that these TPE genomes were virtually identical, demonstrating they share a recent common ancestor and perhaps a single chain of transmission. Finally, we applied deep amplicon sequencing to search for minority *T. pallidum* resistance alleles, showing that RFLP-tested macrolide sensitive samples do not harbour resistant TPE 23S minority alleles, as well as providing the evidence supporting *de novo* evolution of the TPE A2059G mutation within a single patient or lesion.

Our study provides new scientific data about the characteristics, epidemiology, and transmission dynamics of yaws. The genomic dissection renders compelling, high-resolution evidence on the factors that underpin yaws re-emergence and drug resistance after a single round of MDA. These data will support improvement of policies and strategies to eradicate yaws, whilst also informing strategy for MDA campaigns in other diseases where population data is sparse. As part of the global eradication initiative, MDA campaigns against yaws have the potential to significantly reduce transmission of infection in many high-prevalence areas. In these settings, flexible and urgent solutions are needed for a rational approach to counteract recrudescence of infection and drug-resistance. We recommend high MDA coverage to reduce the number of missed people with active or latent yaws (e.g. non-co-operation, working elsewhere, etc), and post-MDA surveillance for the detection of “the last yaws cases” and to control importation and onward transmission of new infections. We also recommend careful monitoring of affected communities during MDA follow-up in order to rapidly detect new emergence of azithromycin resistance, and consideration for using alternative treatments for cases detected post-MDA. Genomic data can play a central role in monitoring and anticipating the dynamics of transmission of yaws to minimize the risk of re-emergence.

## Data Availability

Raw sequencing reads from WGS were deposited at the European Nucleotide Archive (ENA)
under BioProject PRJEB34799. Raw sequencing reads from deep amplicon sequencing were
deposited at the Short Read Archive (SRA) under BioProject PRJNA575636. All accessions used
in this project are listed in Supplementary Data 1 and Supplementary Data 2 along with the
appropriate sample identifier.

https://figshare.com/articles/A_genomic_epidemiology_investigation_of_yaws_re-emergence_and_bacterial_drug_resistance_selection/11418012

## Acknowledgements

This work was funded by Wellcome (Grant #206194) and the Provintial Deputation of Barcelona (Grant #1940000465). The funders played no role in study design, sample collection, analysis or reporting. The authors thank the sequencing team at Wellcome Sanger Institute.

## Figures & Supplementary

**Supplementary Figure 1.**
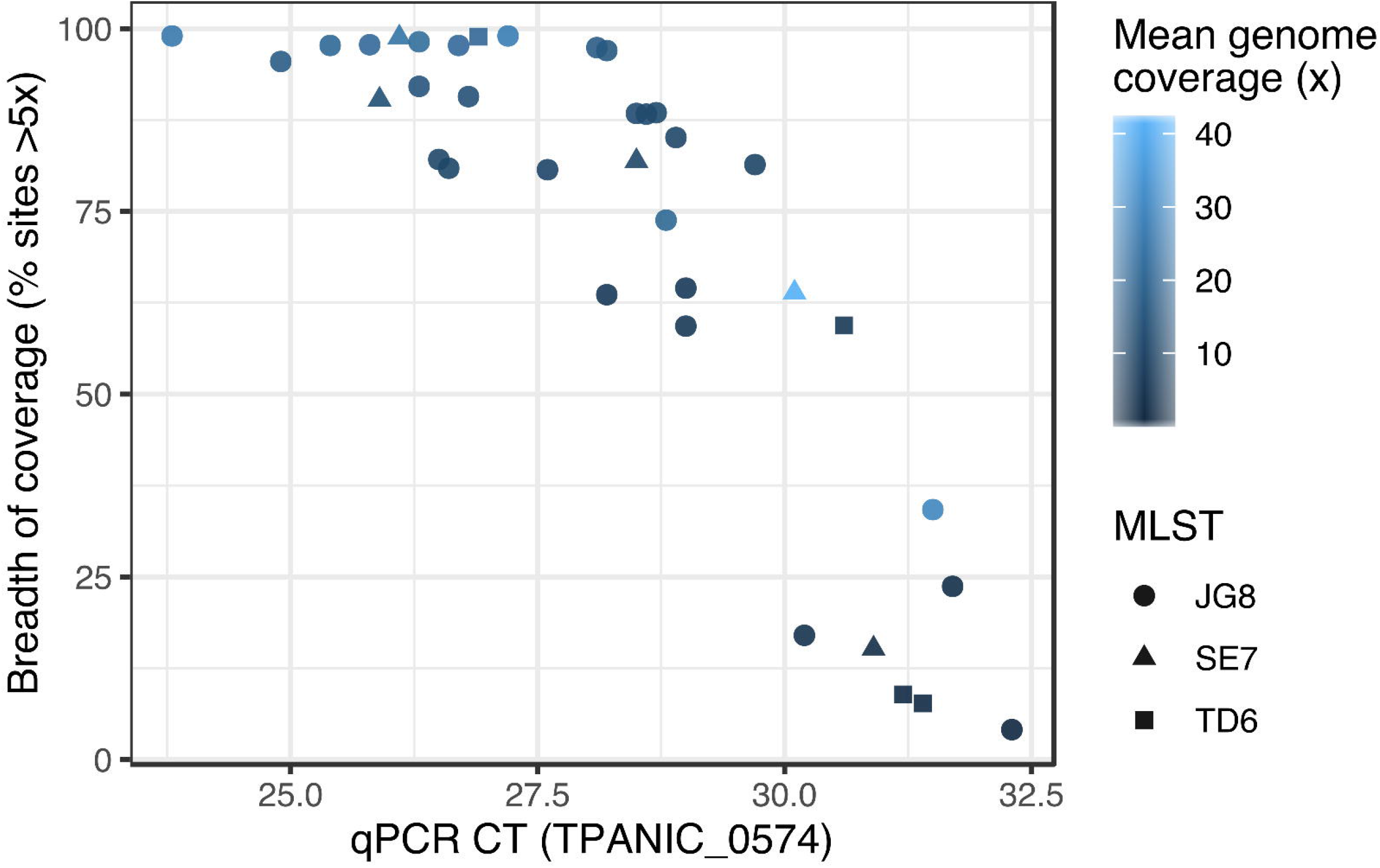
Proportion of whole genome recovery (breadth) according to qPCR Ct. Also showing mean genome coverage (colour) and MLST type (shape).

**Supplementary Figure 2.**
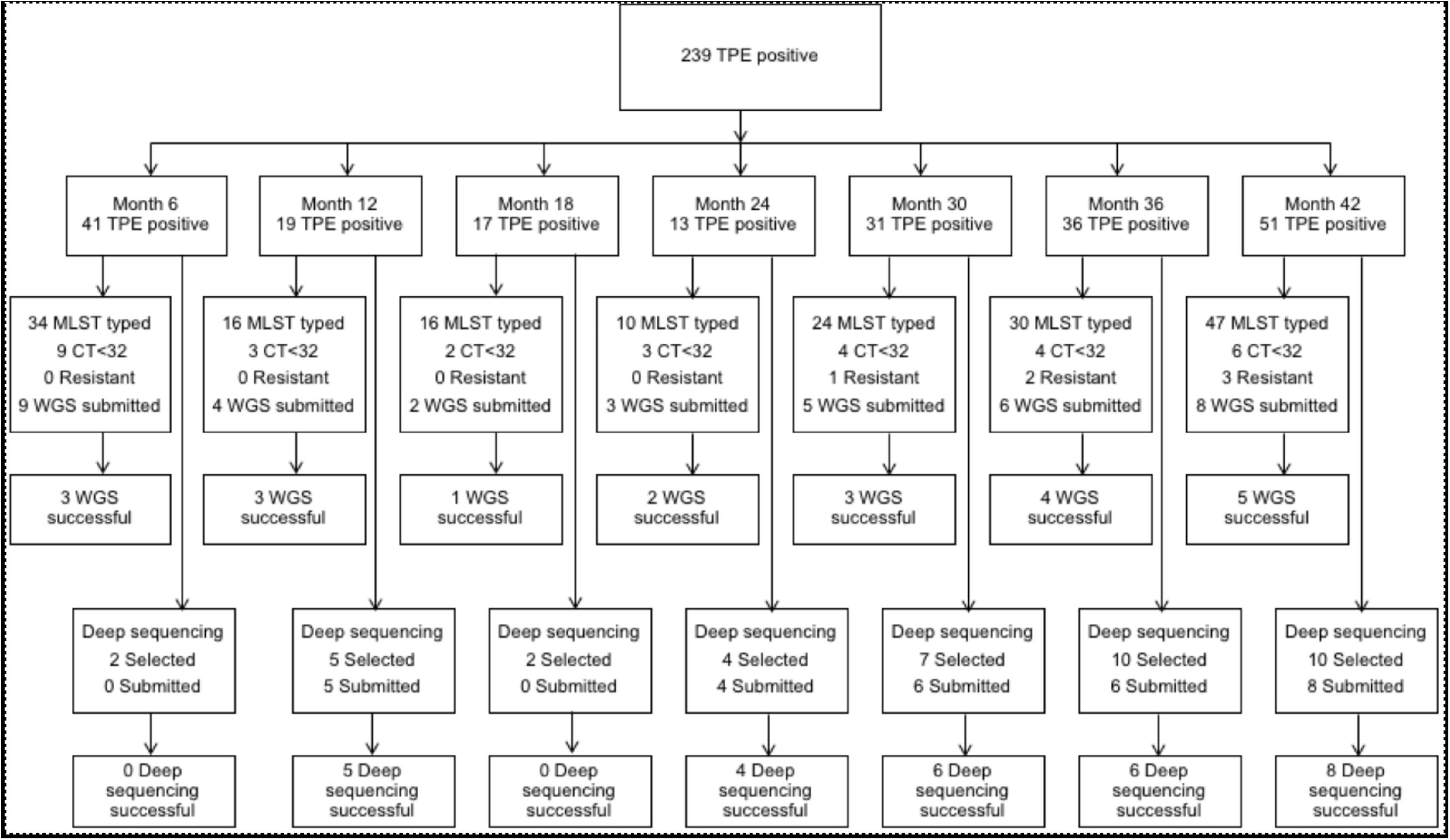
CONSORT diagram of study sample workflows. Samples submitted for WGS were selected to represent MLST types from those with Ct <32, apart from six samples involved in macrolide resistance. Samples submitted for deep sequencing were a temporally selected but otherwise random sample of TPE positive samples, apart from six samples involved in macrolide resistance.

**Supplementary Figure 3.**
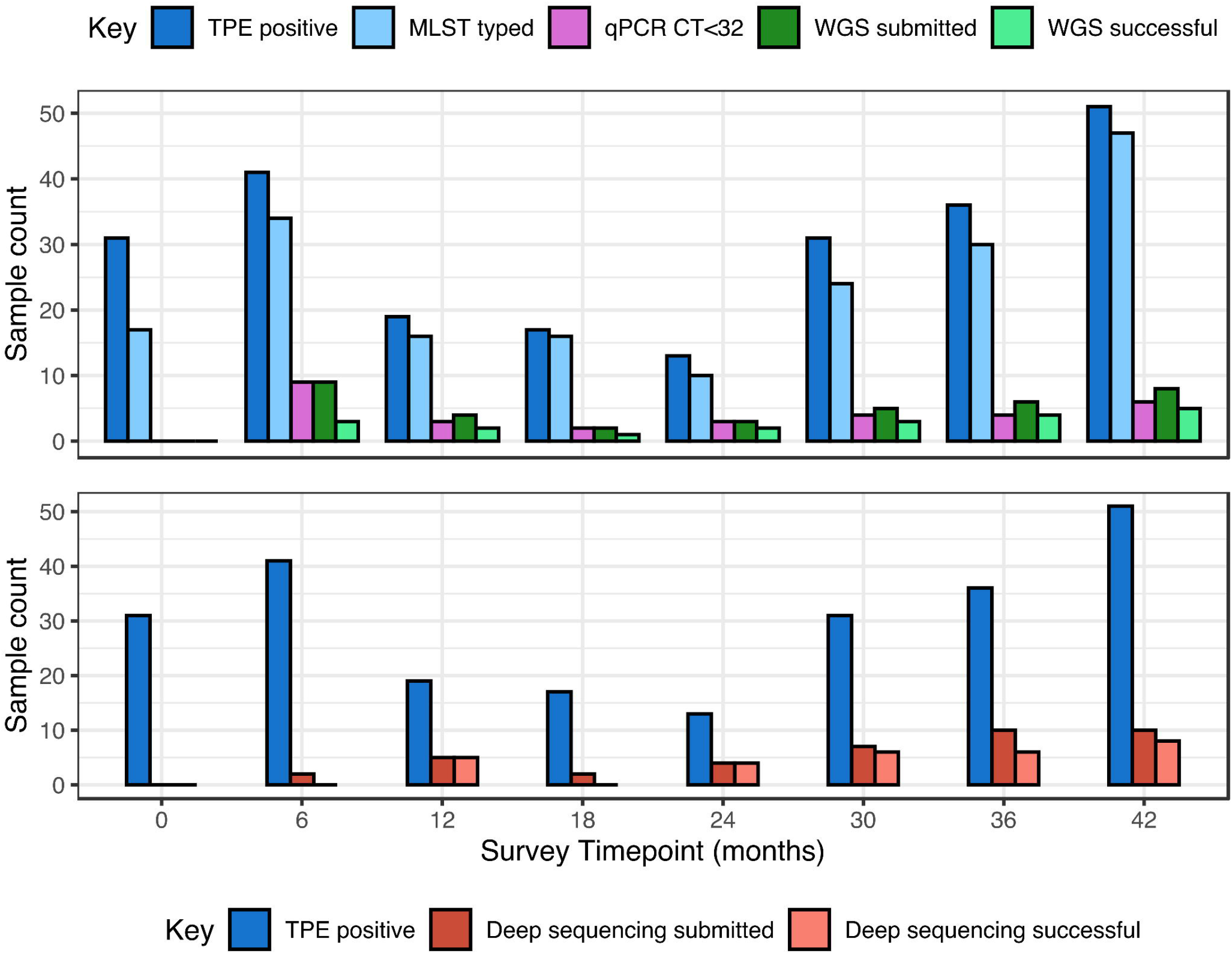
Study sample distribution by survey round for the WGS and deep sequencing workflows. Samples submitted for WGS were selected to represent MLST types from those with Ct <32, apart from six samples involved in macrolide resistance. Samples submitted for deep sequencing were a temporally selected but otherwise random sample of TPE positive samples, apart from six samples involved in macrolide resistance.

**Supplementary Figure 4.**
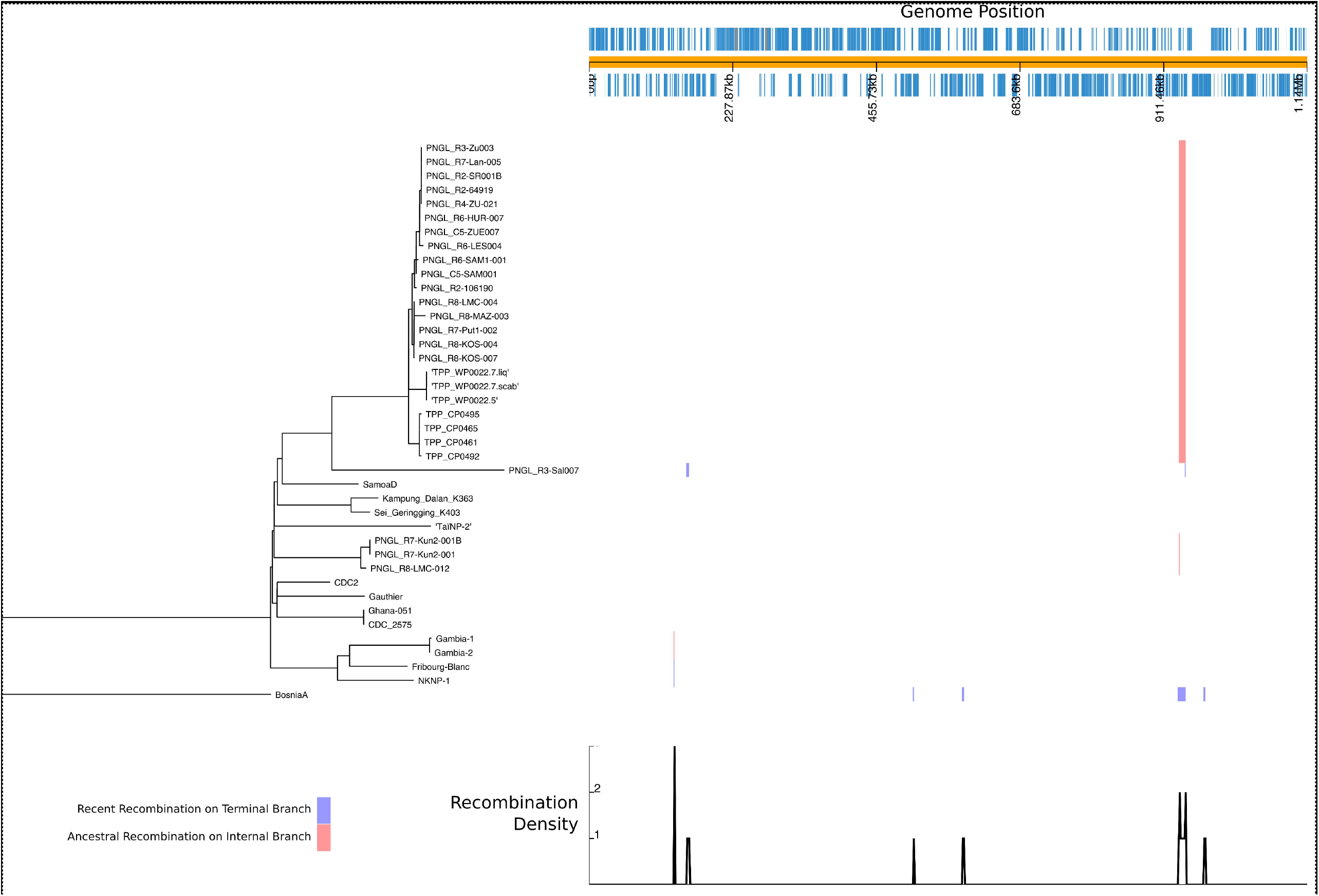
Maximum likelihood phylogeny and recombination analysis of 40 whole TPE genomes (with TPEN BosniaA as outgroup). Plot shows ML phylogeny and analysis of recombining regions identified by gubbins (graph and positional blocks). Red blocks indicate ancestral recombination, while blue blocks indicate recent terminal branch-specific recombination events. The large red block of recombination between 936,491 and 947,537 bp was previously reported in genomes from the Solomon Islands, and is common to JG8 genomes from Papua New Guinea.

**Supplementary Figure 5.**
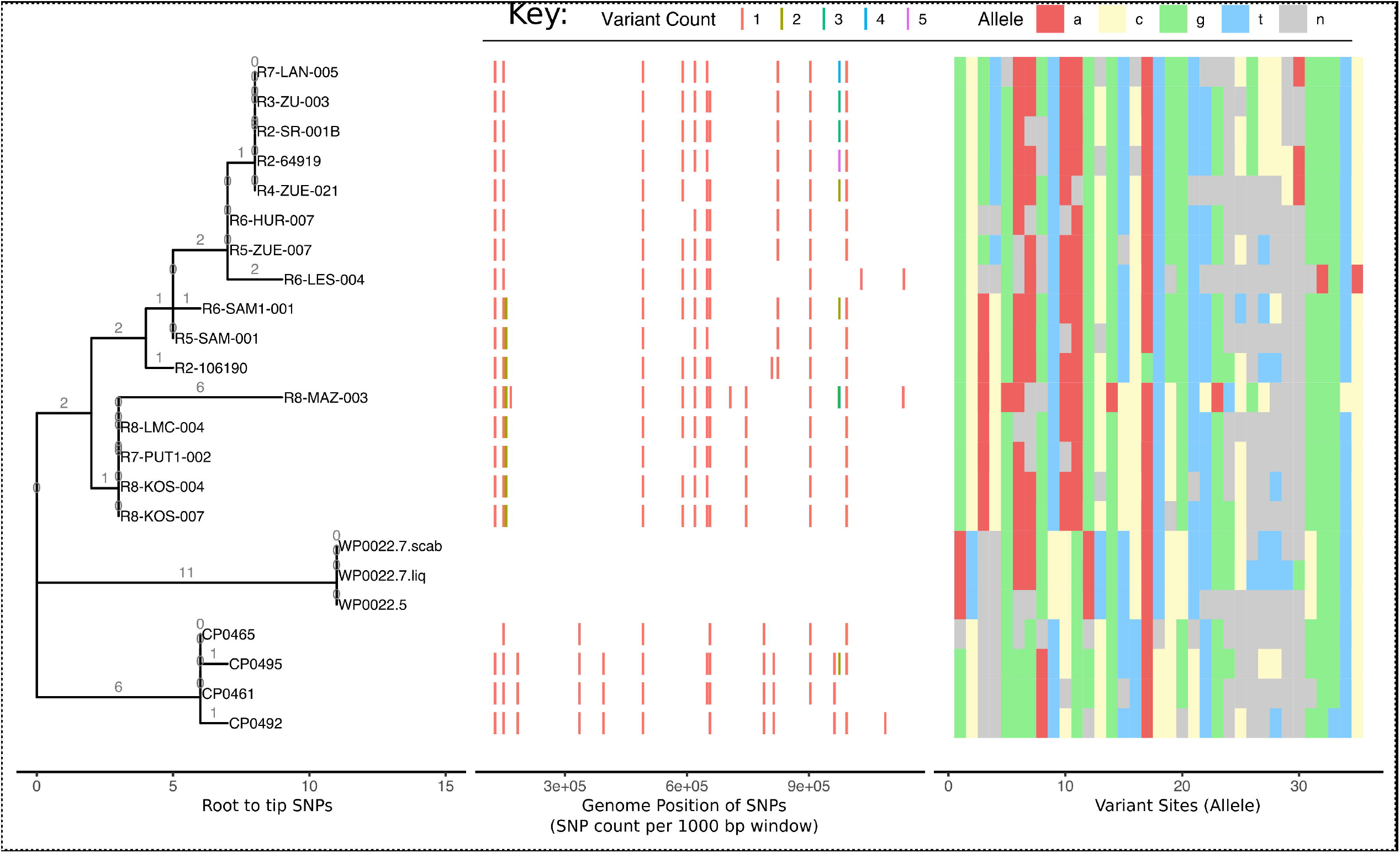
JG8 subtree phylogeny scaled by SNPs, showing distribution of SNPs relative to WP0022.7 genomes, and alleles at each variant site. For some genomes (e.g. CP0465) there was insufficient coverage at certain sites to conclusively determine presence/absence of SNPs; this did not affect the phylogenetic placement of genomes.

**Supplementary Figure 6.**
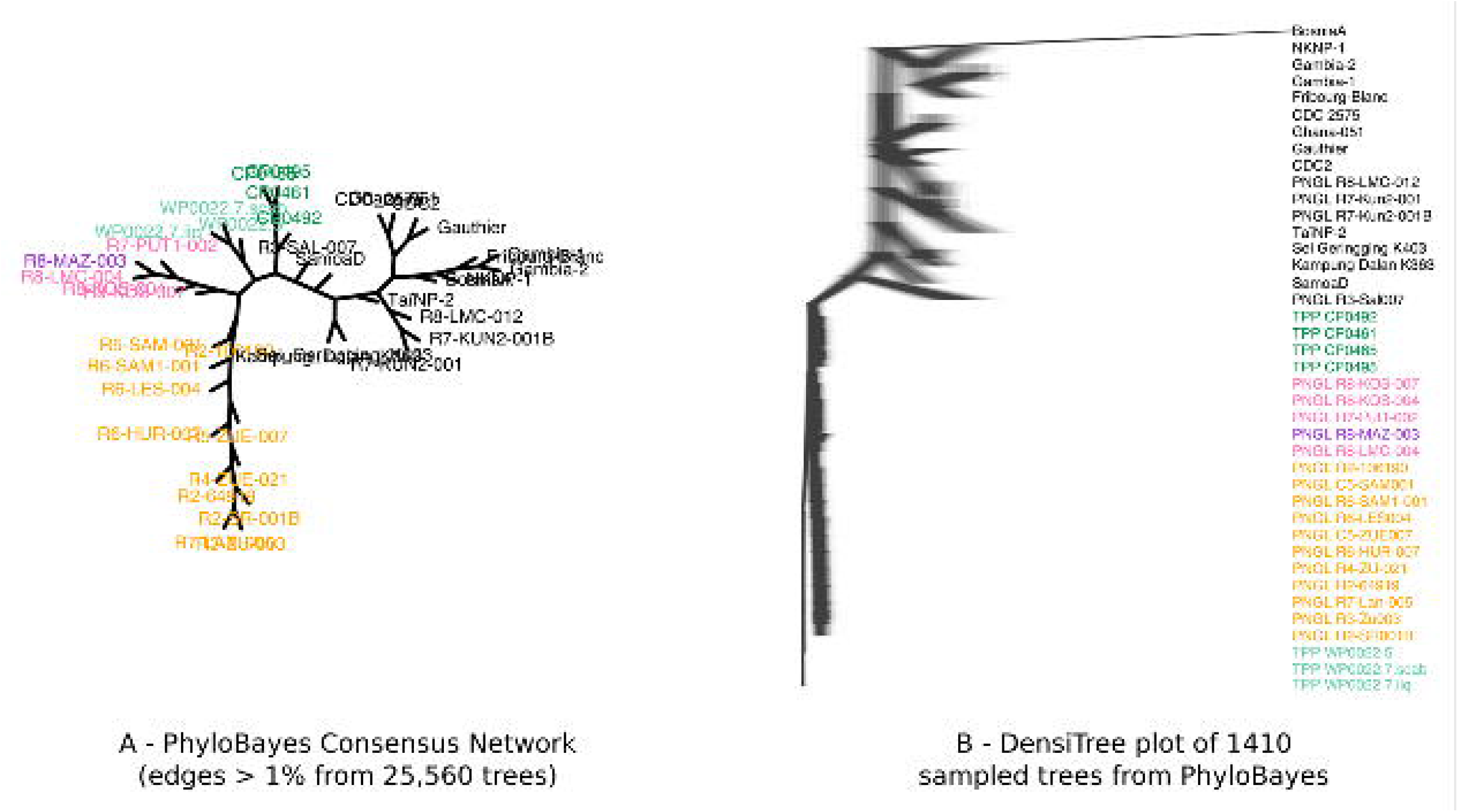
Bayesian analysis supports phylogenetic structure of Maximum Likelihood analyses, including separation of sub-lineages. A – Consensus network constructed from 25,560 PhyloBayes trees, showing no uncertain nodes >1% in the dataset. B – DensiTree plot of 1410 subsampled PhyloBayes trees, reproducing clustering of samples, including JG8 sub-lineages (coloured).

## Supplementary Methods File

**Supplementary Data 1.** Metadata for samples from WGS study

**Supplementary Data 2.** Sequencing read count data for amplicon deep sequencing

